# Common and rare variant analyses implicate late-infancy cerebellar development and immune genes in ADHD

**DOI:** 10.1101/2023.11.30.23299200

**Authors:** Yuanxin Zhong, Larry W. Baum, Justin D. Tubbs, Rui Ye, Lu Hua Chen, Tian Wu, Se-Fong Hung, Chun-Pan Tang, Ting-Pong Ho, Robert Moyzis, James Swanson, Chi-Chiu Lee, Pak C. Sham, Patrick W. L. Leung

**Author notes:** Corresponding authors: Chi Chiu Lee, Kwai Chung Hospital, Hong Kong SAR, China,; Pak Chung Sham, Department of Psychiatry, The University of Hong Kong, Hong Kong SAR, China,; Patrick W.L. Leung, Department of Psychology, The Chinese University of Hong Kong, Hong Kong SAR, China,.

## Abstract

**Background:** Attention-deficit hyperactivity disorder (ADHD) is a common neuropsychiatric disorder with a significant genetic component, characterized by persistent symptoms of inattention, hyperactivity, and/or impulsivity. The latest genome-wide association study (GWAS) meta-analysis of ADHD identified 27 whole-genome significant risk loci in the European population. However, genetic risk factors for ADHD are less well-characterized in the Asian population, especially for rare variants.

**Methods:** Here, we present an analysis of common and rare variant contributions to ADHD in a Hong Kong sample comprising 279 cases and 432 controls, who were genotyped using the Illumina Infinium Global Screening Array.

**Results:** We identified 41 potential genomic risk loci with a suggestive association (*p* < 1e^−4^), pointing to 111 candidate risk genes, which were enriched for genes differentially expressed during late infancy brain development. Furthermore, tissue enrichment analysis implicated the involvement of the cerebellum. *POC1B*, a gene previously found in a genome-wide significant locus of ADHD in the European population, was replicated in the current study, potentially implicating a trans-ancestral effect in ADHD. In addition, an accumulation of ADHD common-variant risks found in European ancestry samples was found to be significantly associated with ADHD in the current study. At the polygenic level, we also discovered a strong genetic correlation with resting-state functional MRI connectivity of the cerebellum involved in the attention/central executive and subcortical-cerebellum networks, which is consistent with the neural pathophysiology for ADHD. In rare variant analyses, we discovered that ADHD cases carried an elevated load of rare damaging variants in *TEP1*, *MTMR10*, *DBH*, *TBCC,* and *ANO1*. ADHD genetic risk was associated with immune processes, demonstrated in both common and rare variant analyses.

**Conclusions:** These findings re-validate the abnormal development of the neural system in ADHD and extend the existing neuro-dysfunction hypothesis to a multi-system perspective.

## Introduction

Attention-deficit/hyperactivity disorder (ADHD) is a neurodevelopmental disorder that has a relatively high prevalence worldwide, around 5% in children and 2.5% in adults (Faraone et al., 2015). In Hong Kong, one in every 15 children has ADHD (Liu, Xu, Yan, & Tong, 2018). It is characterized by inattention, hyperactivity and impulsiveness, with downstream long-term effects on school and work performance, domestic and social relationships, and mental health (Babinski, Neely, Ba, & Liu, 2020; Boomsma, van Beijsterveldt, Odintsova, Neale, & Dolan, 2021; Capusan, Bendtsen, Marteinsdottir, & Larsson, 2019; Jangmo et al., 2021), causing a considerable burden on the family and society (Le et al., 2014; Libutzki et al., 2019; Zhao et al., 2019).

Genetic factors contribute to ADHD, as shown by twin heritability estimates of up to 0.74 (Faraone & Larsson, 2019). The latest large-scale genome-wide association study (GWAS) meta-analysis of ADHD, comprising 38,691 individuals with ADHD and 186,843 controls, revealed 27 genome-wide significant risk loci and highlighted 76 candidate risk genes, which showed enrichment among genes expressed in early brain development (Demontis et al., 2023). This study not only provided deeper insight into the genetic structure of ADHD, but also demonstrated the importance of common variants in ADHD risk accounting for 14-22% of the phenotypic variation (Demontis et al., 2023), consistent with the estimates from previous studies (Lee et al., 2013). However, all cohorts included in this meta-analysis are of European ancestry (Demontis et al., 2023). A previous GWAS with 1,040 ADHD cases and 963 controls, recruited from a homogeneous Han Chinese sample, demonstrated a statistically significant (*p* = 0.0072) but modest (r_g_ = 0.39) correlation in SNP effect sizes with those estimated from the European Ancestry ADHD samples (Yang et al., 2013). These results suggest that there may be substantial heterogeneity in the effects of common genetic variants across Asian and European ancestries, so that larger studies in Asian populations are necessary for understanding their biological mechanisms and producing more accurate risk predictions.

Rare variants, which can contribute up to one-fourth of heritability (Demontis et al., 2019; Lee et al., 2013), are another major genetic contributor to ADHD. ADHD cases display a higher load of rare protein-truncating variants (rPTVs) in genes with high genetic constraint scores (Rajagopal et al., 2022; Satterstrom et al., 2019). A large-scale European-ancestry study suggests a convergence of genes containing common and rare risk variants, by showing that ADHD cases have an excess burden of rPTVs in genes identified from common variant analyses (Demontis et al., 2023). However, due to methodological limitations and the higher cost of whole genome or whole exome sequencing, the analyses of rare variant contributions to ADHD are still in their early stage.

Compared to studies conducted in European populations, ADHD genetic studies in Chinese are still limited in sample size (Huang et al., 2019; Luo et al., 2020; Yang et al., 2013). To better understand the genetic architecture underpinning ADHD in the Chinese population, we tested the contribution of common variants and the burden of rare deleterious variants with samples recruited in Hong Kong. We integrated functional genomics data to identify potentially causal genes and gain biological insights. We characterized the polygenic architecture of ADHD and its overlap with other phenotypes by polygenic score (PGS) analyses.

## Methods

Figure 1 shows an overview of the current study design.

**Figure 1.**
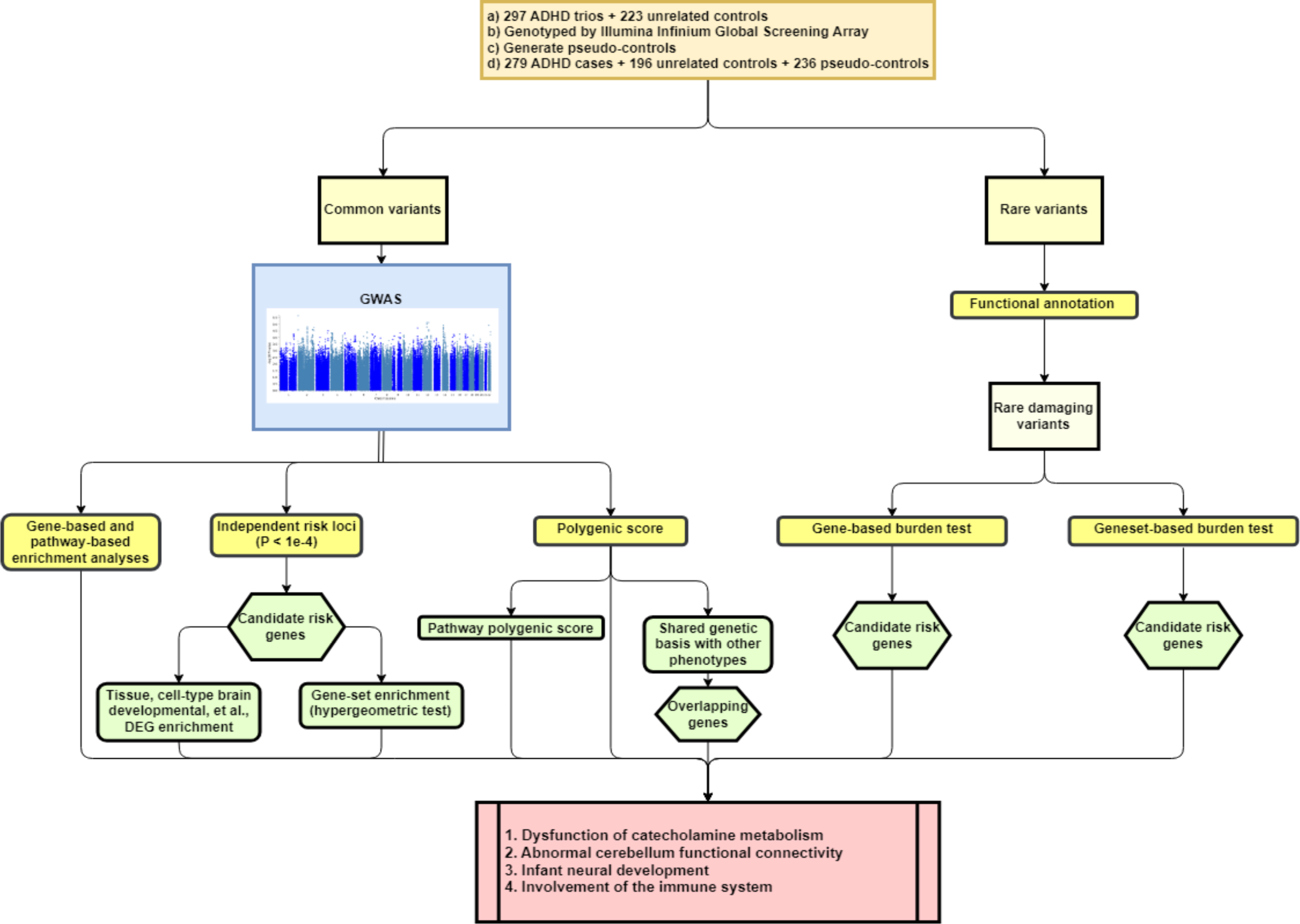
An overview of the current study design. DEG, differentially expressed gene.

### Samples

Participants in this study consisted of 297 boys aged 6–11 years with a primary clinical diagnosis of ADHD (including 254 family trios comprising a boy with ADHD and his two biological parents), and 223 boys without ADHD who served as age- and gender-matched typically-developing controls. Additional inclusion criteria were Chinese ethnicity and studying in local mainstream primary schools. Exclusion criteria for the probands were IQ below 80, psychosis, bipolar disorder, Tourette’s syndrome, chronic motor or vocal tic disorders, or serious neurological or medical conditions. For the controls, the parents were asked a set of screening questions about any behavioural and emotional problems of their children. Children whose parents reported concerns, including any symptoms of ADHD, ASD, or other particular learning issues, or who had received complaints or assessment referrals on these issues from schools or other parties, or who were diagnosed with a psychiatric disorder, were excluded from participation as controls. The probands were recruited from child and adolescent psychiatric services of three local public hospitals, while the controls were recruited from mainstream schools and advertisements in social media.

The Joint Chinese University of Hong Kong-New Territories East Cluster, the Hospital Authority Kowloon Central and Kowloon West Cluster Clinical Research Ethics Committee provided ethical approval for this study, which complies with the most recent Declaration of Helsinki. All participants provided informed consent for the current study. Further details on the constitution and recruitment of the samples can be found from a previous publication (Ma et al., 2021).

### Genotyping, quality control and imputation

Genotyping was performed using the Illumina Infinium Global Screening Array (GSA), whose backbone comprises common and rare variants selected according to research value. Pre-imputation quality control (QC) was performed using established protocols (Marees et al., 2018; Turner et al., 2011). Briefly, we eliminated subjects and SNPs in the analysis according to the following quality control criteria: SNP call rate < 0.98, subject call rate < 0.98, autosomal heterozygosity deviation (|F_het_| > 0.2), difference in SNP missingness rate between cases and controls > 0.02, and deviation from SNP Hardy-Weinberg equilibrium (HWE) (*p* < 1e^−6^). Variants with minor allele frequency (MAF) significantly deviating (*p* < 1e^−3^) from that in the 1000 Genomes Project phase 3 data of the East Asian population (Auton et al., 2015) were also excluded. Based on principal component analysis projecting to the 1,000 Genomes Project phase 3 data of the East Asian population, we removed genetic outliers. For trio data, we generated pseudo-controls from phased haplotypes before imputation. Due to the design features of the GSA, we separated the common (MAF > 0.05) and rare variants according to the above external reference panel (Figure S1).

Imputation of common variants was performed based on 1000 Genomes Project phase 3 data of the East Asian population (Auton et al., 2015) using the Michigan Imputation Server (Das et al., 2016). Variants with imputation INFO score > 0.8 and MAF > 0.05 were included for further analysis of common variants. For rare variants, imputation was not attempted, and we used posterior probabilities (GP) index > 0.8 in the VCF (Variant Call Format) file to ensure the relatively high quality of the variants. Functional annotation was conducted by Variant Effect Predictor (VEP) (McLaren et al., 2016) based on all available implemented data. Only variants with MAF < 0.05 in the 1000 Genomes Project phase 3 data of East Asians (Auton et al., 2015) or in gnomAD (Karczewski et al., 2020), and categorized as having medium or high functional impact by Ensembl, were included in subsequent rare variant analyses.

### Common variant GWAS

GWAS was carried out with the imputed additive genotypes using logistic regression of ADHD cases versus unrelated controls and pseudo-controls in PLINK 1.9 (www.cog-genomics.org/plink/1.9/) (Chang et al., 2015). To correct for population stratification, the top five principal components were added as covariates. A two-sided genome-wide significant P-value threshold of 5e^−8^ and a suggestive P-value threshold of 1e^−4^ (Demontis et al., 2023; Yang et al., 2013) were adopted to allow for multiple testing.

### Gene-based association and gene-set analysis

Gene-based association analysis and gene-set analysis were conducted by MAGMA v.1.08 with default settings (de Leeuw, Mooij, Heskes, & Posthuma, 2015). Gene-based analysis was performed for 19,093 protein-coding genes and we considered the 21 candidate risk genes reported in a previous ADHD GWAS analysis conducted using Chinese Han samples (Yang et al., 2013) separately. For gene-set analysis, testing was done on curated gene sets and Gene Ontology (GO) terms (N = 15,485) from MsigDB (de Leeuw et al., 2015; Liberzon et al., 2011). Five principal components were used as covariates. We used a Bonferroni correction to adjust for multiple testing. Accounting for the number of terms (gene/gene-set) examined, thresholds for genome-wide significance were determined (*p* = 0.05/19,093 = 2.62 e^−6^ for gene-based association analysis, *p* = 0.05/21 = 0.0024 for candidate gene-based association analysis and *p* = 0.05/15,485 = 3.23e^−6^ for gene-set analysis).

### Tissue-specific and cell type-specific gene expression analysis

Correlations between ADHD genome-wide gene-based associations and gene expression patterns in specific tissues were investigated by MAGMA Tissue Expression analysis implemented in the FUMA (Functional Mapping and Annotation) GWAS platform (Watanabe, Taskesen, van Bochoven, & Posthuma, 2017) with expression data from GTEx (54 tissue types) and BrainSpan (brain samples at 29 different ages). Cell type-specific expression analysis was performed by FUMA’s CellType model with 13 single-cell RNA-sequencing datasets from the human brain, the same as those used in a recent ADHD meta-analysis (Demontis et al., 2023).

### Mapping risk variants to genes and enrichment analyses

In addition to gene-based tests on previously reported candidate genes, we also performed further analysis to link risk variants to their potentially novel target genes. We input the GWAS results into FUMA (Watanabe et al., 2017), with pre-defined top-lead SNPs being identified as those with p-values below the suggestive threshold after clumping by PLINK (www.cog-genomics.org/plink/1.9/) (Chang et al., 2015) (r^2^ < 0.1). For each pre-defined top-lead SNP, other SNPs in high linkage disequilibrium (LD) (r^2^ > 0.8) with it were regarded as candidate causal variants that constitute an independent association signal. Then, genomic risk loci were defined from these independent signals by merging those separated by less than 250 kilobases.

In each locus, candidate causal variants were connected to genes based on genomic location, eQTL (expression quantitative trait loci) data, and chromatin interaction mapping data in human brain tissues and blood samples (see Supporting Information), as implemented in FUMA under the default settings (Watanabe et al., 2017). The mapped candidate risk genes were utilized in further gene-set enrichment analyses by FUMA’s GENE2FUNC module to determine whether they were enriched among (1) genes differentially expressed in specific tissues derived from GTEx (Lonsdale et al., 2013) (54 tissue types), (2) genes differentially expressed at specific brain developmental stages derived from BrainSpan (Miller et al., 2014; Sunkin et al., 2012) (brain samples from individuals at 29 different ages and at 11 general developmental stages), and (3) genes encoding proteins in specific pre-defined gene sets implemented in GENE2FUNC. A hypergeometric test was used to determine the statistical significance of the enrichment of ADHD candidate risk genes against all protein-coding genes as background genes. Bonferroni correction was applied to adjust for the multiple testing.

### Polygenic score analysis

We used PRSice2 (Choi & O’Reilly, 2019) to perform polygenic score (PGS) analysis with p-value thresholds (5e^−8^, 1e^−7^, 1e^−6^, 1e^−5^, 1e^−4^, 1e^−3^, 0.01, 0.02, 0.03, 0.04, 0.05, 0.1, 0.2, 0.3, 0.4, 0.5, 1) and an external European LD reference panel generated from the 1000 Genomes Project phase 3 data (Auton et al., 2015). The latest publicly available Psychiatric Genomics Consortium (PGC) summary statistics of five major psychiatric disorders [i.e., ADHD (Demontis et al., 2023), autism spectrum disorder (Grove et al., 2019), bipolar disorder (Mullins et al., 2021), major depressive disorder (Giannakopoulou et al., 2021), and schizophrenia (Trubetskoy et al., 2022)] were used as the training datasets to calculate individual-level PGS in our sample. Regression analyses were performed to evaluate the prediction of case-control status by individual PGS covarying for the first 5 principal components to allow for possible population stratification. Statistical significance was determined by a permutation-based procedure to correct for the testing of multiple p-value thresholds, as implemented by PRSice. For PGS showing nominally significant prediction (empirical *p* < 0.05 after 10000-replicate permutation test), we further conducted a pathway-based PGS analysis for each of the 7,763 GO BP terms from MsigDB (de Leeuw et al., 2015; Liberzon et al., 2011) by PRset with a permutation procedure to take pathway size into account (Choi et al., 2023).

We further calculated PGS for brain-imaging phenotypes using UK Biobank GWAS summary statistics (Elliott et al., 2018) to identify brain regions or connections related to ADHD. In order to reduce the number of brain-imaging phenotypes, we used only those related to the cerebellum based on results from the tissue-specific enrichment analysis described above. Thus, 72 structural MRI features and 260 resting-state fMRI features related to the cerebellum were included in this analysis. Individual PGSs were calculated by PRSice2 as above, except that UK Biobank data were used to derive the LD reference panel. Bonferroni correction for testing 332 phenotypes was applied to the permutation p-values (significance *p*-value threshold = 0.05/332 = 1.51e^−4^).

### Identifying overlapping risk loci and genes

ADHD risk loci and genes that overlap with other phenotypes were identified by two analyses: (1) Phenome-Wide Association Studies (PheWAS) for the suggestive GWAS signals in the current study for associations with a wide range of other traits. Top-lead SNPs and their high LD proxies (r^2^ > 0.8) were used to search the GWAS Catalog (Sollis et al., 2022) and PhenoScanner v2 (Kamat et al., 2019) for associations (*p* < 5e^−8^); (2) For psychiatric or brain-imaging traits whose PGS models significantly predicted case-control status in our ADHD sample, we conducted variant-to-gene mapping analyses (as described above) on their GWAS summary statistics to identify risk variants and mapped candidate genes for these psychiatric or brain-imaging traits in FUMA (Watanabe et al., 2017). Genes overlapping with the detected ADHD candidate risk genes were further analyzed for their tissue-specific expression profiles and biological functions.

We estimated the correlation in genetic effects between the large European ancestry ADHD GWAS (the latest from PGC ADHD) and the Chinese ADHD sample in the current study using Popcorn, which is designed to manage trans-ethnic scenarios (Brown, Ye, Price, & Zaitlen, 2016). We also conducted genetic correlation analyses between ADHD and the psychiatric or brain-imaging phenotypes whose PGS models predict ADHD, using GWAS summary statistics from European samples and LD score regression (Bulik-Sullivan et al., 2015).

### Burden testing of rare variants

Gene-based and gene set-based association analyses on our ADHD sample were carried out using the SKAT test (Wu et al., 2011) implemented in RVTESTS (Zhan, Hu, Li, Abecasis, & Liu, 2016). We first performed functional annotation for all rare variants (MAF < 0.05) using Ensembl Variant Effect Predictor (VEP) (McLaren et al., 2016). Then, we extracted the rare variants predicted to have high or medium damaging effects on protein function and identified their canonical transcripts and annotated genes. All genes with >2 damaging rare variants across ADHD and control samples (N = 4800) were included for gene-based association testing. For the gene set-based association test, we defined gene sets according to GO BP terms from MsigDB (de Leeuw et al., 2015; Liberzon et al., 2011) (N = 7763). Five genetic principal components were used as covariates in both gene-based and geneset-based tests. Since the genes/genesets are correlated, we performed a 10000-time permutation test implemented in the program for testing significance, which preserved the family-wise error rate (FWER) without over-correction (Purcell et al., 2014), and *p*_FWER_ < 1e^−3^ was regarded as significant.

## Results

### Genetic association analyses and correlations with gene expression

The GWAS analysis identified 48 independent lead variants (r^2^ < 0.1) located in 41 genomic loci at the suggestive threshold (*p* < 1e^−4^) (see Figure 2 and Table 1). None of them reached the genome-wide significance threshold (*p* < 5e^−8^). Gene-based analysis did not show a significant association after multiple testing adjustments. Of the previously identified candidate genes (Yang et al., 2013), the most significant was *KCTD16* (*p* = 0.0086), which did not survive Bonferroni adjustment for 21 tests (Table S1). There was no significant result from the gene-set based association analyses after multiple testing adjustments.

**Figure 2.**
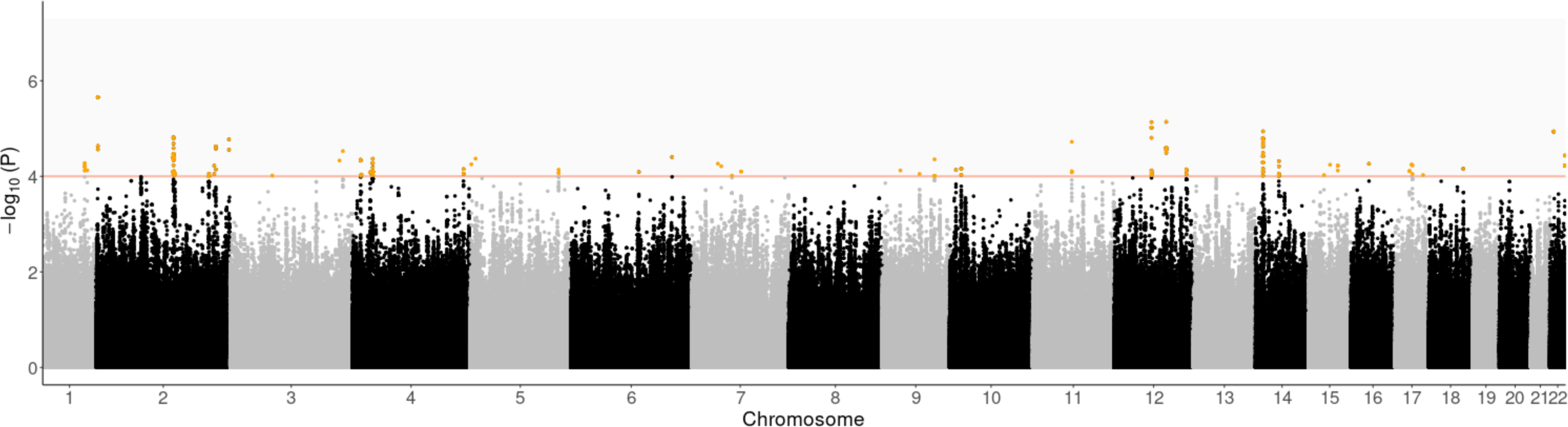
Manhattan plot of GWAS of 279 ADHD cases and 432 controls from Hong Kong samples. Orange line shows the suggestive threshold (*p* = 1e^−4^) adopted in the current study. Identified genomic loci are highlighted in orange (N = 41).

**Table 1.** Results for the 41 index variants identified at the suggestive threshold in the GWAS of 279 ADHD cases and 432 controls.

| Genomic Locus | Chr | Position (bp) | rsID | Effect/<br>Non-effect | MAF | OR | SE | P value | NearestGene |
| --- | --- | --- | --- | --- | --- | --- | --- | --- | --- |
| 1 | 1 | 208968342 | rs17013389 | A/C | 0.199 | 0.540 | 0.152 | 5.27E-05 | RP11-459K23.2 |
| 2 | 1 | 218106816 | rs6667062 | C/G | 0.431 | 1.578 | 0.115 | 7.48E-05 | LINC00210 |
| 3 | 2 | 1623553 | rs72763534 | A/G | 0.200 | 1.994 | 0.146 | 2.21E-06 | AC144450.1 |
| 4 | 2 | 139917014 | rs12613166 | T/A | 0.334 | 0.594 | 0.120 | 1.53E-05 | AC062021.1 |
| 5 | 2 | 141478468 | rs436323 | T/C | 0.058 | 2.651 | 0.248 | 8.59E-05 | LRP1B |
| 6 | 2 | 207843712 | rs1006389 | C/T | 0.099 | 0.456 | 0.202 | 9.92E-05 | CPO |
| 7 | 2 | 218862241 | rs1001098 | T/C | 0.431 | 1.541 | 0.110 | 8.98E-05 | TNS1 |
| 8 | 2 | 221965082 | rs6739103 | A/G | 0.272 | 0.560 | 0.137 | 2.35E-05 | AC011233.2 |
| 9 | 2 | 241754034 | rs73012754 | A/G | 0.259 | 1.736 | 0.128 | 1.68E-05 | KIF1A |
| 10 | 3 | 64173901 | rs1035275 | C/T | 0.236 | 0.580 | 0.140 | 9.65E-05 | PRICKLE2:PRICKLE2-AS3 |
| 11 | 3 | 184176114 | rs9854433 | C/A | 0.495 | 1.597 | 0.112 | 2.96E-05 | EIF2B5 |
| 12 | 4 | 12889878 | rs223917 | C/T | 0.205 | 1.841 | 0.150 | 4.49E-05 | RP11-22A3.2 |
| 13 | 4 | 14744974 | rs10010400 | C/T | 0.071 | 2.601 | 0.244 | 9.20E-05 | LINC00504 |
| 14 | 4 | 28040589 | rs6840350 | A/G | 0.190 | 0.525 | 0.164 | 8.28E-05 | AC007106.1 |
| 15 | 4 | 31290243 | rs60139026 | G/T | 0.453 | 1.588 | 0.113 | 4.29E-05 | RP11-315A17.1 |
| 16 | 4 | 183085906 | rs11935992 | G/A | 0.119 | 0.432 | 0.211 | 7.06E-05 | TENM3:RP11-402C9.1 |
| 17 | 5 | 159955315 | rs1895317 | T/C | 0.207 | 0.561 | 0.146 | 7.28E-05 | ATP10B |
| 18 | 6 | 90023021 | rs79488521 | A/T | 0.385 | 0.631 | 0.117 | 8.08E-05 | GABRR2 |
| 19 | 6 | 144268094 | rs6930726 | A/C | 0.314 | 1.649 | 0.122 | 3.96E-05 | PLAGL1 |
| 20 | 7 | 37427783 | rs60704414 | G/A | 0.326 | 1.660 | 0.126 | 6.14E-05 | ELMO1 |
| 21 | 7 | 53186040 | rs6954000 | T/G | 0.465 | 0.639 | 0.115 | 9.67E-05 | HAUS6P1 |
| 22 | 7 | 71145041 | rs376468499 | T/C | 0.141 | 1.793 | 0.148 | 7.93E-05 | WBSCR17 |
| 23 | 9 | 90691910 | rs12684075 | T/C | 0.491 | 1.546 | 0.111 | 8.95E-05 | RP11-350E12.4 |
| 24 | 9 | 112592855 | rs7867595 | A/G | 0.140 | 1.866 | 0.153 | 4.41E-05 | PALM2:PALM2-AKAP2:AKAP2 |
| 25 | 10 | 8109576 | rs3847417 | T/G | 0.282 | 1.594 | 0.118 | 7.34E-05 | GATA3 |
| 26 | 10 | 15117423 | rs12219813 | A/G | 0.359 | 0.619 | 0.120 | 6.94E-05 | DCLRE1CP1 |
| 27 | 11 | 70405685 | rs7127137 | C/T | 0.496 | 0.642 | 0.112 | 7.82E-05 | SHANK2 |
| 28 | 12 | 62006325 | rs4758832 | T/C | 0.184 | 0.516 | 0.148 | 7.33E-06 | RP11-410B16.1 |
| 29 | 12 | 90095173 | rs11105379 | C/T | 0.210 | 1.853 | 0.137 | 7.26E-06 | ATP2B1 |
| 30 | 12 | 124361292 | rs56044291 | T/C | 0.054 | 2.741 | 0.254 | 7.15E-05 | DNAH10 |
| 31 | 14 | 33554821 | rs6571581 | A/C | 0.497 | 0.613 | 0.112 | 1.15E-05 | NPAS3 |
| 32 | 14 | 58228104 | rs1950997 | A/G | 0.442 | 0.615 | 0.120 | 4.79E-05 | SLC35F4 |
| 33 | 15 | 55136295 | rs1915202 | C/T | 0.194 | 1.635 | 0.126 | 9.44E-05 | RP11-548M13.1 |
| 34 | 15 | 81166740 | rs28384338 | A/G | 0.394 | 1.566 | 0.112 | 5.98E-05 | KIAA1199 |
| 35 | 16 | 49946670 | rs6500265 | T/C | 0.217 | 1.686 | 0.129 | 5.46E-05 | RP11-305A4.3 |
| 36 | 17 | 34350579 | rs7208039 | T/C | 0.162 | 0.518 | 0.167 | 7.77E-05 | CCL23 |
| 37 | 17 | 40582296 | rs1905339 | C/T | 0.350 | 0.618 | 0.119 | 5.70E-05 | PTRF |
| 38 | 17 | 43313404 | rs9891552 | A/T | 0.063 | 2.589 | 0.237 | 5.92E-05 | FMNL1 |
| 39 | 18 | 65038142 | rs12605748 | G/T | 0.187 | 1.808 | 0.149 | 6.93E-05 | RP11-563H6.1 |
| 40 | 22 | 26915245 | rs8135081 | A/G | 0.171 | 1.996 | 0.158 | 1.16E-05 | CTA-445C9.15 |
| 41 | 22 | 49817197 | rs7287788 | A/G | 0.119 | 0.453 | 0.192 | 3.67E-05 | C22orf34 |

The location [chromosome (Chr.) position (bp)] maps in hg19. MAF shows frequency of effect allele in cases and controls. OR of the effect with respect to effect allele. NearestGene, the gene nearest the SNP based on ANNOVAR annotations implemented by FUMA with default settings.

Gene-based association test statistics for protein-coding genes showed a nominally significant correlation with gene expression levels in the caudate nucleus (beta = 0.015, SE = 0.0083, *p* = 0.036) and the nucleus accumbens (beta = 0.025, SE = 0.0080, *p* = 0.037) of the basal ganglia. Cell-type-specific gene-expression levels did not show a significant correlation with ADHD gene-based association results.

### Risk variant-to-gene mapping and enrichment analyses

We identified 111 ADHD candidate risk genes (Table S2) from mapping based on the genomic locations, eQTLs and chromatin interactions of the 48 independent lead variants and their high-LD neighboring SNPs. These candidate genes included *POC1B*, which was reported in a recent large-scale PGC ADHD meta-analysis (Demontis et al., 2023), and *SHANK2*, which was described in our previous study (Ma et al., 2021).

Bi-clustering analysis of gene expression data (across 111 genes and 53 tissues) revealed a cluster of 16 genes with over expression in brain tissues generally and a smaller cluster of 3 genes with over expression specifically in the cerebellum (Figure S2). In the further enrichment analyses, we found that these 111 candidate risk genes were significantly enriched among up-regulated genes in the brain at 1 year postnatal (*p*_unadj_ = 5.42e^−4^, *p*_adj_ = 0.016) or at the late-infancy development stage, which is defined as between 6 and 12 months postnatal (*p*_unadj_ = 3.06e^−5^, *p*_adj_ = 3.37e^−4^). These genes were also significantly enriched among up-regulated genes in the cerebellar hemisphere (*p*_unadj_ = 7.35e^−4^, *p*_adj_ = 0.039) and cerebellum (*p*_unadj_ = 7.79e^−4^, *p*_adj_ = 0.041), and enriched at a nominally significant level among differentially expressed genes in the substantia nigra (*p*_unadj_ = 0.0038, *p*_adj_ = 0.20) (Figure 3). They were also highly enriched in a GO cellular components term, presynaptic active zone (*p*_unadj_ = 4.35e^−5^, *p*_adj_ = 0.044); reported genes in GWAS of response to cognitive-behavioral therapy in anxiety disorder (*p*_unadj_ = 2.29e^−6^, *p*_adj_ = 0.0021); and several Reactome immune signaling pathways, including interleukin 9 signaling (*p*_unadj_ = 1.36e^−5^, *p*_adj_ = 0.013), interleukin 21 signaling (*p*_unadj_ = 1.94e^−5^, *p*_adj_ = 0.013), signaling by leptin (*p*_unadj_ = 2.66e^−5^, *p*_adj_ = 0.013), interleukin 15 signaling (*p*_unadj_ = 5.79e^−5^, *p*_adj_ = 0.022) and signaling by cytosolic fgfr1 fusion mutants (*p*_unadj_ = 1.28e^−4^, *p*_adj_ = 0.038) (Figure S3).

**Figure 3.**
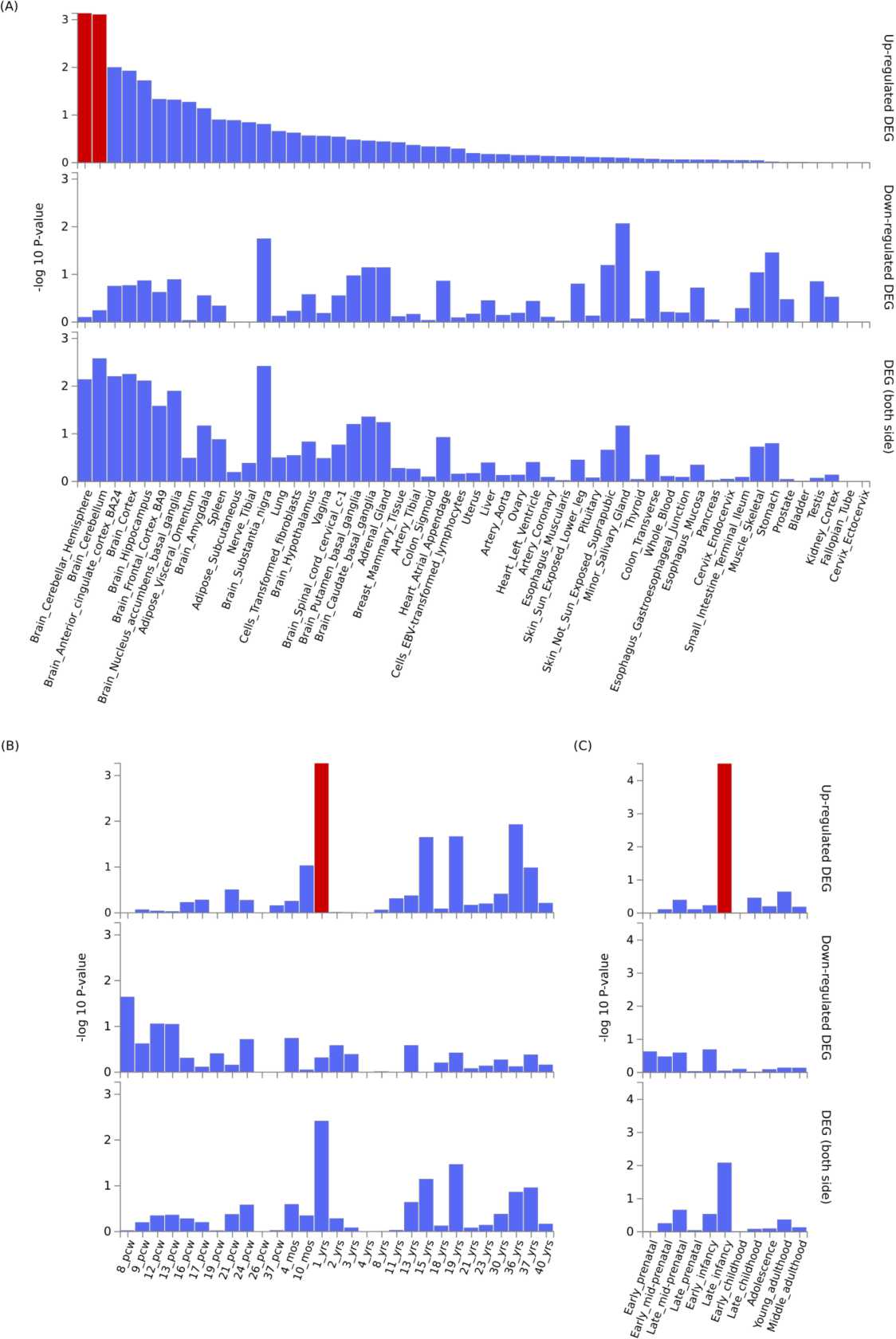
Enrichment in 111 ADHD candidate risk genes among differentially expressed gene (DEG) sets across tissues and in brain tissue. (A) Enrichment among DEGs across 53 specific tissues types available in the GTEx v.7 database. Enrichment was also evaluated among DEGs in brain tissue from Brain Span representing (B) 29 different specific ages and (C) 11 general developmental stages. Highlighted red bars indicate significant enrichment at Bonferroni corrected *p* ≤ 0.05.

### Polygenic score analysis

Among the PGSs for five major psychiatric disorders, only the ADHD PGS significantly predicted ADHD case-control status in the current GWAS after permutation tests (*p*-value threshold = 0.2, Lee.R^2^ = 0.012, empirical-*p* = 0.024). Further pathway-based PGS analysis with PGC ADHD summary statistics found that, among all GO terms, “central nervous system projection neuron axonogenesis” achieved the lowest p-value after 10000-replicate set permutations (Lee.R^2^ = 0.027, competitive-*p* = 1.0e^−4^). Among the PGSs for cerebellum-related brain-imaging phenotypes, the PGS for resting-state fMRI (rs-fMRI) connectivity between two components derived from independent component analysis, [Parietal|Frontal] and [Cerebellum|Temporal], which are related to attention/central executive and subcortical-cerebellum networks, had the best performance and passed Bonferroni correction (Lee.R^2^ = 0.027, empirical-*p* = 1.0e^−4^) (Figure 4).

**Figure 4.**
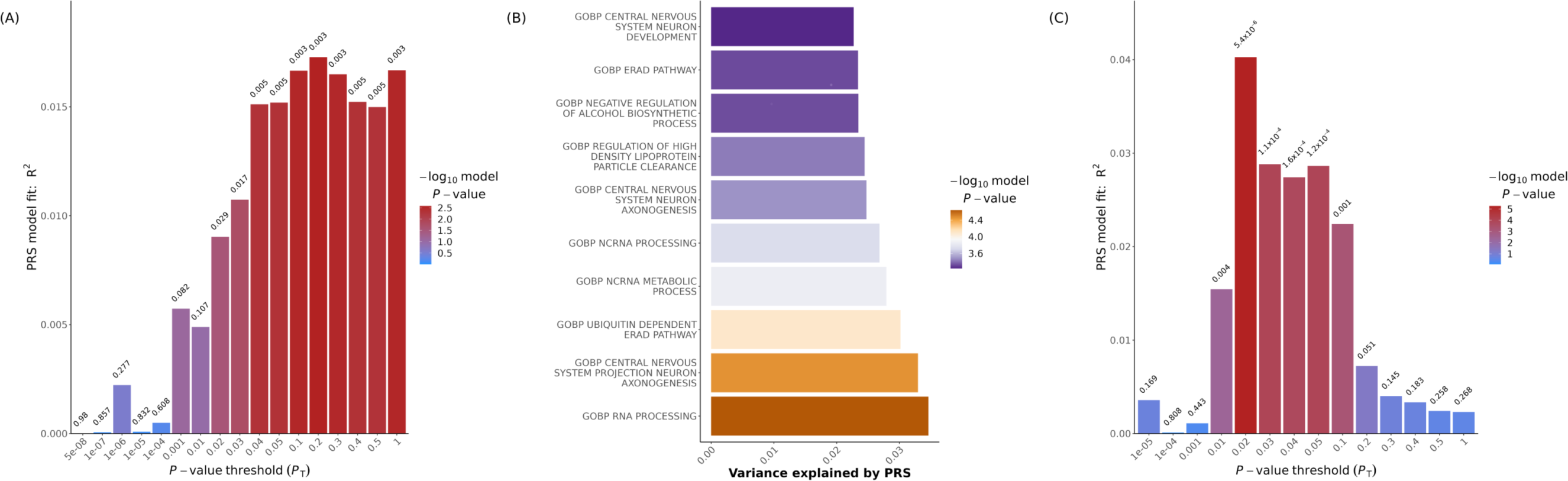
Polygenic risk score (PRS) for ADHD and brain-imaging phenotype. (A) Bar plot of R^2^ for ADHD prediction across multiple *p*-value thresholds in PGC ADHD summary statistics as the training set. (B) Bar plot shows the enrichment of ADHD signals in different biological pathways. (C) Bar plot of PRS for resting-state fMRI (rs-fMRI) connectivity between [Parietal|Frontal] and [Cerebellum|Temporal] showing the explained variance for ADHD at multiple *p*-value thresholds in the Hong Kong sample.

### Genetic overlap of ADHD with other phenotypes

Among top-lead SNPs and their high LD proxies (r^2^ > 0.8), there were significant associations in GWAS of other phenotypes (details in the Table S3). However, there was no enrichment of these phenotypes in any specific phenotype categories defined in PhenoScanner (Kamat et al., 2019). We found two shared genes between our 111 ADHD candidate genes and the risk genes indicated by the rs-fMRI connectivity GWAS data, *LRP1B* and *TENM3*, which have an enhanced expression level in the brain and inhibitory neurons (Karlsson et al., 2021; Uhlén et al., 2015).

There was no significant trans-ancestry genetic correlation between the European ancestry ADHD GWAS and the current ADHD GWAS, however, our power to detect such correlation is low due to the limited sample size. We did not find a significant genetic association of the European ancestry ADHD with rs-fMRI connectivity between [Parietal|Frontal] and [Cerebellum|Temporal] components.

### Rare variant burden test

The gene-based rare variant association test revealed five significant genes, *TEP1* (*p*_FWER_ = 1.58e^−4^), *MTMR10* (*p*_FWER_ = 3.24e^−4^), *TBCC* (*p*_FWER_ = 3.78e^−4^), *DBH* (*p*_FWER_ = 4.31e^−4^), and *ANO1* (*p*_FWER_ = 8.08e^−4^), after 10000-replicate permutation test. In the gene-set-based rare variant analysis with 5860 biological process GO terms, “norepinephrine biosynthetic process” (*p*_FWER_ = 3.30e^−4^) reached significance.

## Discussion

Although our study was underpowered to identify genome-wide significant signals for ADHD, a substantial proportion of the 41 independent risk loci identified as passing the suggestive threshold (*p* < 1e^−4^) are likely to be truly associated with ADHD. These loci implicated 111 candidate risk genes through functional genomic data. The genes were significantly enriched among genes up-regulated in the cerebellum compared to other tissues. The traditional view of the cerebellum is that it plays a critical role in the fine control and coordination of movements to achieve automatic and smooth execution with practice (Manto et al., 2012; Paulin, 1993). Growing evidence suggests that the cerebellum is similarly involved in the fine control and coordination of perceptual, cognitive and emotional processes (Baumann & Mattingley, 2012; Buckner, 2013; Deverett, Kislin, Tank, & Wang, 2019; Mariën & Borgatti, 2018).The symptoms of ADHD may partly reflect the dysregulation of attention, impulses, emotional responses, and motor activities, resulting from cerebellar dysfunction (Castellanos, Sonuga-Barke, Milham, & Tannock, 2006; Coghill, Toplak, Rhodes, & Adamo, 2018; Goetz et al., 2017). Consistent with a role of the cerebellum in ADHD, reduced cerebellar volume has been reported in ADHD (Wyciszkiewicz, Pawlak, & Krawiec, 2017) and to be associated with greater symptom severity (Castellanos et al., 2002).

Using polygenic scores, we found ADHD to be genetically correlated with the resting-state functional MRI connectivity between the attention/central executive and subcortical-cerebellum networks. Disordered connectivities of the cerebellum with the frontal cortex and basal ganglia have significant influences on attention, working memory, planning, inhibition, and coordinated movements, required for flexible, adaptive behaviors (Clark, Semmel, Aleksonis, Steinberg, & King, 2021; Sathyanesan et al., 2019). The disruption of these processes has been proposed as being fundamental in the genesis of ADHD symptoms (Bush, 2010; Oldehinkel et al., 2016; Stoodley, 2016). The overlapping risk genes between ADHD and this functional connectivity are *LRP1B* and *TENM3*. *LRP1B* is a cell-surface receptor which is expressed in the brain and is involved in cell migration and synaptic plasticity (Príncipe, Dionísio de Sousa, Prazeres, Soares, & Lima, 2021). *LRP1B* was associated with motor and cognitive ability in 2-year old (Sun et al., 2020) and with hyperlocomotion in people using cocaine (Vorspan et al., 2020). *LRP1B* was the only gene found to have altered expression in both ventral and dorsal striatum by repeated administration of cocaine in rats (Vorspan et al., 2020). *TENM3* is a Teneurin involved in guiding cortical neuronal migration and synapse formation (Del Toro et al., 2020) in the hippocampus (Berns, DeNardo, Pederick, & Luo, 2018), thalamus and striatum (Tran, Sawatari, & Leamey, 2015). *TENM3* knockout mice were delayed in developing motor skills (Tran et al., 2015). Teneurins are genetically associated with bipolar disorder and schizophrenia (Yi et al., 2021). *TENM3* is associated with childhood autoimmune diseases (Li et al., 2015), and the presence of autoantibodies to N-methyl-d-aspartate-receptor subunit-NR1 (NMDAR1), the most common antigen for anti-brain autoantibodies (Daguano Gastaldi et al., 2023). These results suggest that some of the identified variants are relevant to ADHD by altering neuronal development and the coordinated functioning of the cerebellum and other brain regions.

The current study provides evidence supporting the importance of the catecholamine system, especially in the cerebellum, in ADHD. We identified the rare-variant associations of ADHD with *DBH* (Dopamine beta-hydroxylase), whose encoded protein converts dopamine to norepinephrine. The identified ADHD candidate genes were nominally enriched among genes expressed in the midbrain dopaminergic regions (Paladini & Tepper, 2016). The primary pharmacologic effect of psychostimulant drugs for ADHD is to elevate the activity of central dopamine and norepinephrine (Faraone, 2018). Interestingly, several studies have demonstrated the crucial role of the cerebellum in dopamine deficit-related neurological and psychiatric disorders (Miquel, Gil-Miravet, & Guarque-Chabrera, 2020; Parker, Narayanan, & Andreasen, 2014; Wu & Hallett, 2013). Recent neuroimaging evidence indicates connectivity between cerebellum and midbrain dopaminergic regions (substantia nigra and ventral tegmental area), both structurally (Bareš et al., 2015; Cacciola et al., 2017; Flace, Livrea, Galletta, Gulisano, & Gennarini, 2020; Milardi et al., 2016) and functionally, and that this connectivity is altered in Parkinson’s disease (O’Shea, Popal, Olson, Murty, & Smith, 2022). In addition, lesional and axonal tracing experiments on rodents have shown that the midbrain dopaminergic cell groups are the origin of the cerebellar extrinsic dopaminergic fibers (Flace et al., 2021; Ikai, Takada, Shinonaga, & Mizuno, 1992). In mouse models, dopamine acts on D2 receptors in parts of the cerebellum to alter the propensity for social novelty and sociability without changing their motor coordination and other functions (Cutando et al., 2022). A direct efferent influence from the cerebellum to the midbrain dopaminergic nuclei to modulate reward processing and social behavior has also been demonstrated (Carta, Chen, Schott, Dorizan, & Khodakhah, 2019). These studies indicate potential reciprocal, direct connectivities between the cerebellum and the midbrain dopaminergic regions, and the alterations in these functional connectivities may affect cognitive and emotional regulation and social behavior and lead to psychiatric symptoms. Cerebellar function is also modulated by norepinephrinergic input from the locus coeruleus as part of its effects on attention, arousal, and cognition (Breton-Provencher, Drummond, & Sur, 2021; Schwarz & Luo, 2015).

The significant enrichment of the candidate risk genes up-regulated in late-infancy/1-year brain development indicates that this period may be critical for ADHD vulnerability. This stage is a landmark for human nervous system development with a rapid increase in synaptic density in the cerebral cortex, and a greater rate of growth of the cerebellum relative to the rest of the brain. The ratio of the volume of the cerebellum to the intracranial volume reaches a plateau in late infancy. Thus, any disruptions to neural processes such as synaptic proliferation, formation, and pruning at this stage may have a disproportionate effect on the cerebellum, and subsequently produce symptoms of cerebellar dysfunction present in ADHD (Clark et al., 2021; Ivanov, Murrough, Bansal, Hao, & Peterson, 2014; Wyciszkiewicz et al., 2017).

The current study demonstrated, through pathway-enrichment and set-based association analyses, convergent risk of common and rare variants that inform potential biological mechanisms in ADHD. An elevated load of rare, damaging variants in *MTMR10*, *DBH, ANO1, TEP1*, and *TBCC* was observed in ADHD patients. According to Genecard (Safran et al., 2021; Stelzer et al., 2016), *MTMR10*, Myotubularin related protein 10, has an enhanced expression level in oligodendrocytes and astrocytes and is part of “Brain - Nervous system development” and “Astrocytes - Nervous system maintenance” annotation clusters. *ANO1*, encodes a protein required for the Ca^2+^-dependent process extension of radial glial cells, especially in the developing brain, and is involved in regulating the excitability of neurons. *TEP1*, Telomerase associated protein 1, belongs to the “B-cells - Humoral immune response” cluster. *TBCC*, Tubulin folding cofactor C, is annotated to “Immune cells - Immune response” and “T-cells - Immune response” expression clusters. These results implicate, in addition to neural processes, the involvement of the immune system in ADHD. There is indeed increasing evidence supporting the association of immune system dysregulation with psychiatric disorders, including schizophrenia (Miller, Buckley, Seabolt, Mellor, & Kirkpatrick, 2011; Potvin et al., 2008; Sekar et al., 2016), depression (Tubbs, Ding, Baum, & Sham, 2020), and autism spectrum disorder (Al-Haddad et al., 2019; Atladóttir et al., 2010; Masi et al., 2015). Previous studies showed that immune processes might regulate the nervous system and have an impact on the elimination and plasticity of synapses during development (Dantzer, 2018). It has been proposed that modern lifestyles (e.g., hygiene) might have deprived infants of co-evolved immunoregulatory organisms, with adverse effects on neural development and increasing vulnerability to psychiatric disorders (Rook, Lowry, & Raison, 2015).

In the current study, an accumulation of common risk variants for ADHD identified in European ancestry GWAS (measured by a PGS) was found to be significantly associated with case-control status in our Asian ancestry sample. This is consistent with the moderately large estimated SNP-based genetic correlation of 0.39 for ADHD between Europeans and Chinese (Yang et al., 2013). *POC1B*, a gene previously found in a genome-wide significant locus of ADHD in the European population (Demontis et al., 2023), was replicated in the current study, potentially implicating a trans-ethnic effect on ADHD. *KCTD16* showed a nominally significant association with ADHD in a previous Chinese ADHD GWAS study (Yang et al., 2013) and in the current study. These results demonstrate that at least some genetic factors are shared across ancestry groups, supporting the existence of common biological mechanisms underpinning ADHD.

The current study has some limitations. First, the sample size is limited, affecting the statistical power to detect variants with modest effect size and identify individual risk genes. Second, all participants are male, which may affect the generalization of the results. Nevertheless, these current findings likely foreshadow the identification of individual risk genes in larger East Asian cohorts or meta analyses.

## Conclusion

As far as we know, this is the first GWAS study using a Hong Kong ADHD sample. The current study identified convergent risk factors from common and rare variants, which implicates vulnerability in late-infancy brain development, affecting especially the cerebellum, and the involvement of immune processes. Furthermore, our findings show that diverse ancestry groups share some genetic factors driving ADHD.

## Data Availability

All data produced in the present study are available upon reasonable request to the authors

## Conflict of Interest

The authors declare no conflicts of interest.

## Acknowledgements

This work was supported by a General Research Fund (GRF) of the Hong Kong Research Grants Council (RGC) (RGC 449511). Funding sources play no role in study design, data collection, analysis, interpretation, manuscript preparation, or submission.

We are very grateful to the child and adolescent psychiatric services of three public hospitals in Hong Kong, namely, Kwai Chung Hospital, Alice Ho Miu Ling Nethersole Hospital, and United Christian Hospital, which assisted in the recruitment of ADHD children. Special thanks must also be accorded to the hard work of our two former research assistants, Sharon Cheung and Fungyee Ching. Finally, we are very thankful to the children, parents and schools which so willingly participated in this project.

## Notes

### Competing Interest Statement

The authors have declared no competing interest.

### Author Declarations

The Joint Chinese University of Hong Kong-New Territories East Cluster, the Hospital Authority Kowloon Central and Kowloon West Cluster Clinical Research Ethics Committee gave ethical approval for this work.

